# Does Women Empowerment alone Influence Contraception utilization in Bangladesh perspective? Findings from demographic health survey 2017-18 using A SEM analysis

**DOI:** 10.1101/2023.12.05.23299504

**Authors:** Md. Nawal Sarwer, Effat Ara Jahan, Akibul Islam Chowdhury

## Abstract

**Background:** Women empowerment is a crucial issue that is less studied as a factor of contraception use among married women which helps to achieve sustainable development goals (SDGs). The present study aims to assess the relationship between women empowerment and contraception use.

**Methods:** This cross-sectional study used Bangladesh Demographic and Health Survey data 2017-18 which included 12006 (weighted) women aged from 15-49 years old. Hierarchical logistic regression and structural equation model (SEM) were used to show the relationship between women empowerment and contraception use.

**Results:** Overall, the tend to using contraception was increased with increased age, urban residence, increased wealth index and education level of both husband and wife. The findings from regression model showed that women empowerment in terms of women decision making, attitude to violence and social independence significantly influence the contraception use after controlling the covariates (p<0.05). SEM analysis showed negative relationship with overall women empowerment and contraception use (β= -0.138) which was not significant (p>0.05).

**Conclusion:** This study implies that greater women empowerment may not always act as stronger determinant for using contraception, and therefore other contributing factors such as age, education, religion, husband’s participation, joined decision making, economic status and couple relationship should be warranted.

## Introduction

Women’s empowerment has grown in importance on the worldwide development agenda in recent decades which is used as common development strategy in developing countries (1). Access to contraception through family planning programs boosts their chances of having the number of children they want and reduces the chance of unintended pregnancy which lead to abortion related morbidity and mortality (2). According to a recent report about Millenium Development Goals (MDGs) achievement and preparation of Sustainable Development Goals (SDGs) demonstrated that only few countries successfully achieve the target of reducing maternal and child mortality (3). The goal five of SDGs focus on the women empowerment and gender equality.

The idea of “empowerment” for women is broad and covers a variety of topics, including the capacity for decision-making, autonomy, mobility, reproductive rights, economic independence, and legal rights to equal treatment, inheritance, and protection from discrimination. These factors work together to provide women more control over their family planning and reproductive decisions. The need to strengthen efforts to address structural factors that influence the use of contraceptives, such as women’s empowerment, and to better understand these factors is highlighted by behaviors (4). There are direct or indirect connections between women’s empowerment and the use of contraception as a method of managing fertility. Early 1990s data showed that empowered women in rural Bangladesh were more likely to use contraception than other women (5). In recent years, there has been a growing focus on how gender-based power dynamics in men’s and women’s sexual interactions affect attitudes and behaviors connected to fertility (6).

Bangladesh is a Muslim-majority country and men are systematically having more power and authority in decision making process than women (7). However, Bangladesh has made significant progress in increasing the contraceptive prevalence rate (CPR) over the past few decades. The contraceptive prevalence rate (CPR) in Bangladesh increased from 8% in 1975 to 61.9% in 2017 (BDHS-2017) although this prevalence is slightly lower than BDHS 2014 data (62.4%) (8). According to a number of studies, the use of contraception is positively impacted by the empowerment of women in the marital sphere. Women’s empowerment and gender equality frequently play a significant effect in the use of contraceptives and the fall in fertility (9-11). Women’s autonomy significantly affects the usage of contraceptives, especially when it comes to issues involving reproductive rights. When women feel empowered, they are more likely to exercise their right to control their fertility and opt for reliable contraception (4). Women are more likely to have access to and be able to afford various forms of contraception if they are economically independent and actively working. So, increased use of contraception and financial independence are linked (12). A number of previous studies in Bangladesh evaluated the relationships between contraception use and women empowerment which were focused on women’s involvement in decision making (13-17), attitude to violence (15), freedom of movement (15-17). These variables provide a narrow scope of women empowerment for understanding the relationship of women empowerment and contraceptive use.

Therefore, the aim of the present study is to evaluates the relationship between women’s empowerment and the utilization of contraception in Bangladesh using the recent developed index named Survey-based Women’s emPowERment (SWPER) index (18, 19). Calculation of women empowerment is difficult due to its multidimensional term and complex nature. To better capture the multifaceted and complex construct of women’s empowerment and allow for cross-country and cross-temporal comparability Ewer-ling and colleagues created the SWPER index based on individual-level DHS data from 34 African nations and recently it is validated in Bangladesh (18, 19). In Bangladesh, it is crucial to understand whether or not women’s empowerment is now influencing their use of contraception. The study tries to explore the intricate relationship between women’s empowerment and the utilization of contraception in Bangladesh.

## Methods

### Study design

This cross-sectional study was based on the secondary data of the recent Bangladesh Demographic and Health Survey (BDHS) 2017 (8). Data was obtained from the DHS website (https://dhsprogram.com/). The BDHS survey is a nationally representative survey which was conducted by the National Institute for Population Research and Training in collaboration with ICF International and Mitra and Associates. This survey was based on a two-stage stratified sample of households. This survey selected 20,250 households for data collection and collected 20,127 data from ever-married women aged 15-49 covering both rural and urban areas (response rate 99%). The detailed about the survey’s sampling methods, study design and data collection can be found from the following link ((https://dhsprogram.com/methodology/survey/survey-display-536.cfm). The current study included 12006 currently married women who delivered at least one child.

A flow chart of selecting sample for the current study is depicted in Figure 1.

**Figure 1:**
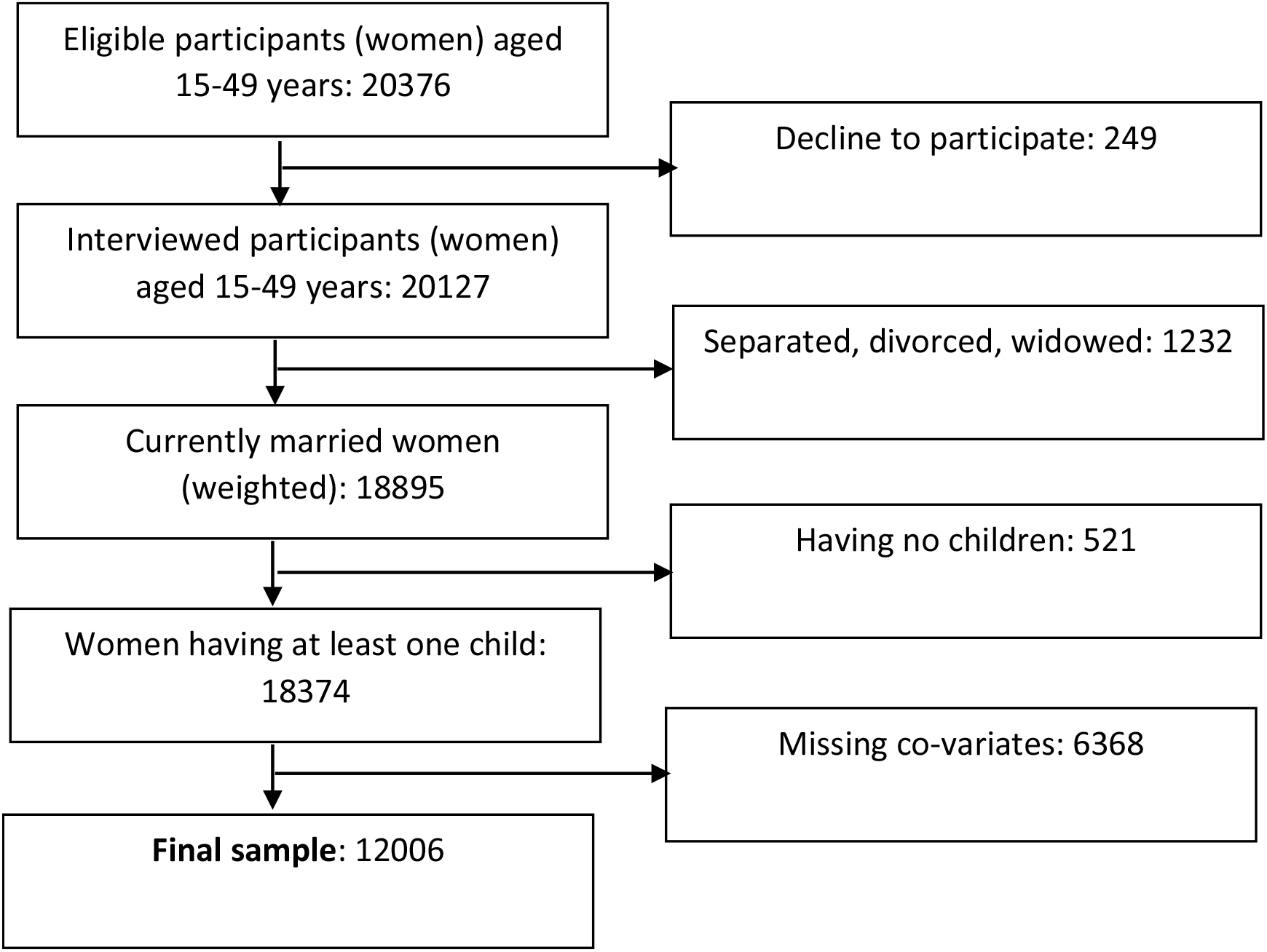
A flowchart of sample selection.

### Exposure, Outcome and covariate variables

The effect of women empowerment on current contraception use was evaluated in the present study. The categories of exposure, outcome and covariate variables were presented in Table 1.

**Table 1:** Description of exposure, outcome and covariate variables.

|  |  |
| --- | --- |
| **Exposure variables** |  |
| Women empowerment | Based on SWPER index, 3 domains |
|  | i. Decision making (low, medium, high empowerment) |
|  | ii. Attitude to violence (low, medium high empowerment) |
|  | iii. Social independence (low, medium high empowerment) |
| **Outcome variables** | Current contraceptive use (Yes, No) |
|  | Methods of contraceptive use (No method, Folkloric method, traditional method, modern method) |
| **Covariates** | Respondent's age (<24 years, 24-34 years, >34 years) |
|  | Education (Primary/no education, secondary, higher) |
|  | Religion (Islam, others) |
|  | Household wealth (Poorest, poorer, middle, richer, richest) |
|  | Area of residence (rural, urban) |
|  | Divisions |
|  | Husband's education (primary/No education, secondary, higher) |
|  | Number of living child (<2, 2-4, >4) |

### Estimation of women empowerment

In the present study, women empowerment was measured using a survey-based women’s empowerment (SWPER) index (18, 19). SWPER index was developed and validated for global (including Bangladesh) which contains three domains: social independence (based on education, age at marriage and first child, and differences in age and education with partners), decision making (question related making decision in the household and to the women’s work) and attitudes to violence (includes five questions about respondent’s opinion on husband beating). In total 14 items were extracted from the DHS survey data to measure the women empowerment and principal component analysis (PCA) was used to construct the index. To measure the index, items were recoded as described in supplementary Table 1 and calculate the standard score of these three domains using the formula of SWPER index. Standard cut-off points were used suggested by the author to categorize the domains of SWPER into low, medium and high empowerment.

### Statistical analysis

Data were recoded, cleaned, and analyzed by using Excel and SPSS software version 23.0. Descriptive statistics were performed to present frequencies and percentages of variables. A Chi-square test was performed to evaluate the correlation of outcome variables with predictors. Multinomial logistic regression model was used to assess the factors affecting the utilization of contraceptive and methods of using contraception. Adjusted odd ratio are presented to assess the strengths of association. A hierarchical logistic regression was also performed where each of variables is entered separately to assess the influence of women empowerment on contraception use.

Structural equation model (SEM) was performed to test the hypothesized relationships of independent and dependent variables. The SEM analysis was performed based on maximum likelihood method and covariance matrix using SPSS Amos 23.0. Fitness of model was estimated using Root Mean Square Error of Approximation (RMSEA), Bentler’s Comparative Fit Index (CFI) and Tucker-Lewis Fit Index (TLI) (20). A p value less than 0.05 is considered as statistically significant.

### Ethical consideration

The study was based on secondary data and data were publicly available in DHS website. Therefore, there is no need further ethical approval. Data accessibility is gained by sending a request to DHS website.

## Results

Table 2 presents the frequency and percentage of variables studied in the present study. There was a slightly equal distribution of respondents among administrative division and residential area. More than 88% of the women were Muslim. The weighted prevalence of higher education among respondents was 20.4% and husband was 25.5%. More than one-third women had 2-4 living children. The wealth index showed that more than half of the women belonged from middle to richest family.

**Table 2:**
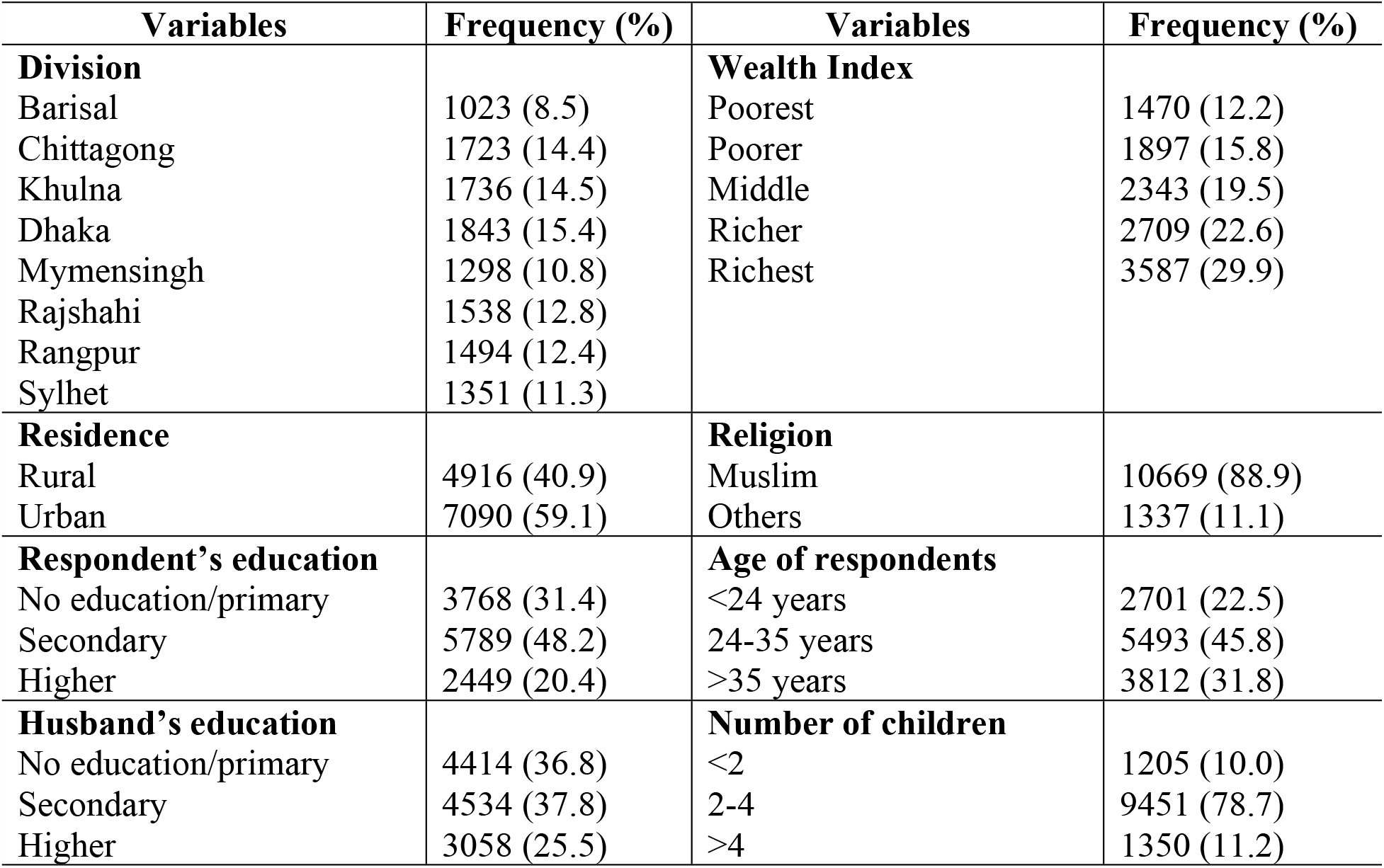
Characteristics of respondents.

Table 3 depicts the percentage distribution of outcome variables in regard to the main exposure variables. Contraception use among women was not found to associate with women’s attitude to violence and social independence. Women empowerment in decision making showed significant association with contraception use as well as with method of contraceptives. More than 90% women who had medium to high decision-making power in family used different types of contraceptive methods.

**Table 3:**
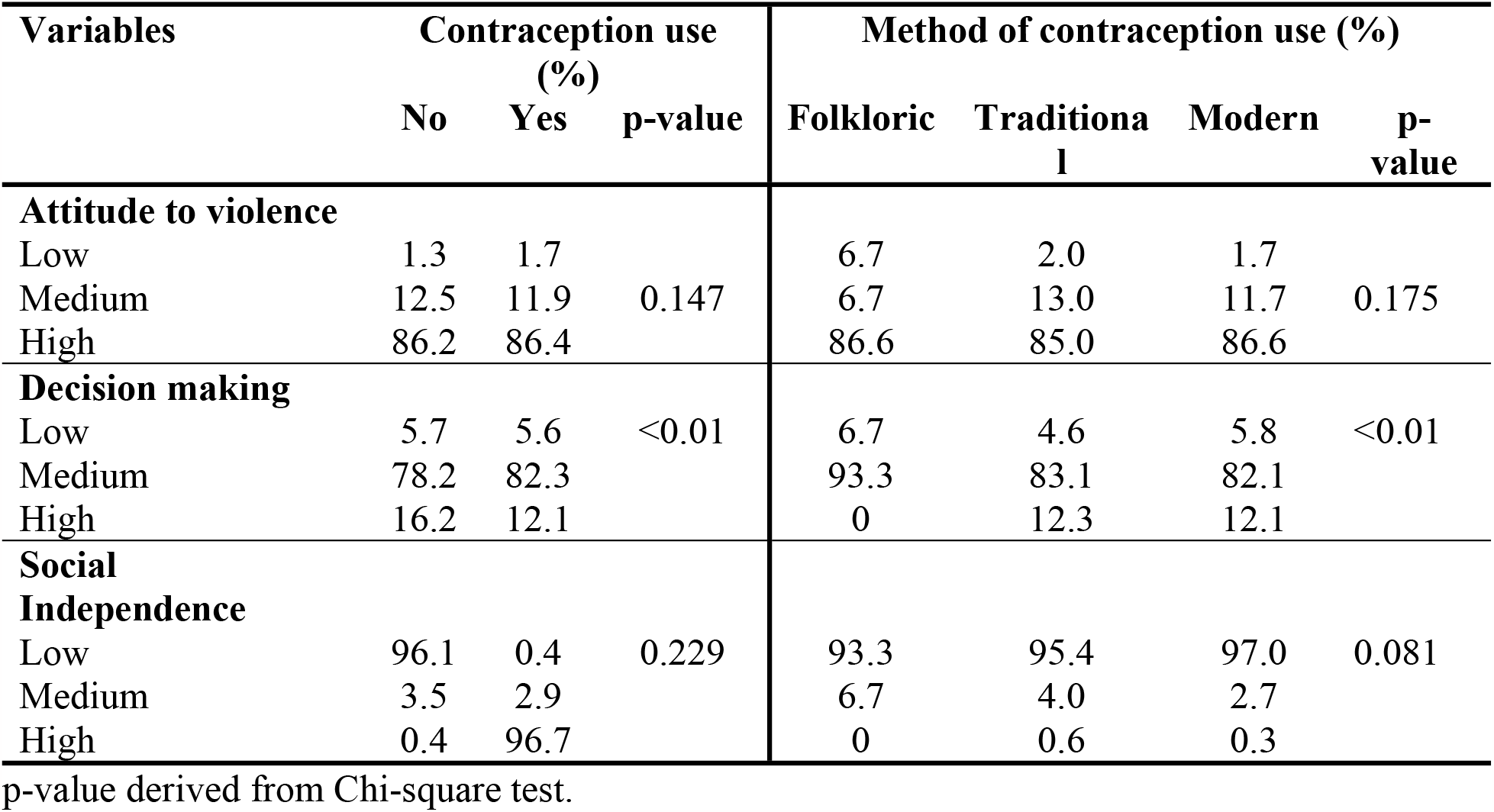
Distribution of outcome variables based on exposure variables.

Women’s age, education and partner’s education showed significant association with contraceptive use and method of contraceptives. Compared with women <24 years of age, women >35 years of age were 1.64 times (AOR: 1.64, 95% CI:1.45-1.85) more likely to use contraceptive and 1.09 times (AOR: 1.09, 95% CI: 1.05-1.14) more likely to use modern contraceptive methods. Highly educated mothers had 1.35 times (AOR: 1.35, 95% CI: 1.15-1.58) higher possibility of use contraceptives compared with women who did not receive any formal education or completed primary education. Area of residence was also showed significant association with contraceptive use and method of contraception use. Muslim women had 0.81 (AOR: 0.81, 95% CI: 0.70-0.93) times likely to use contraception compared with others religion’s women. Women who belonged from richest family, their chance of using contraception and modern contraceptive methods were increased by 0.78 times (AOR: 0.78, 95%CI: 0.65-0.92) and 0.92 times (AOR: 0.92, 95% CI: 0.86-0.97) respectively.

The association of women empowerment in terms of attitude to violence, decision making power, and social independence with contraception use was shown in Table 5 after controlling the covariates (individual, and locality factors). The findings from hierarchical logistic regression showed that women who had medium level attitude to violence and social independence had significantly 0.715 times (AOR: 0.715, 95% CI: 0.515-0.994) and 0.81 times (AOR: 0.81, 95%CI: 0.655-1.002) greater chance to use contraception than women who had low level of attitude to violence and social independence respectively. The odd of using contraception was increased by 0.772-fold (AOR: 0.771, 95% CI: 0.641-0.930) when women had high level of decision-making power in family.

**Table 4:**
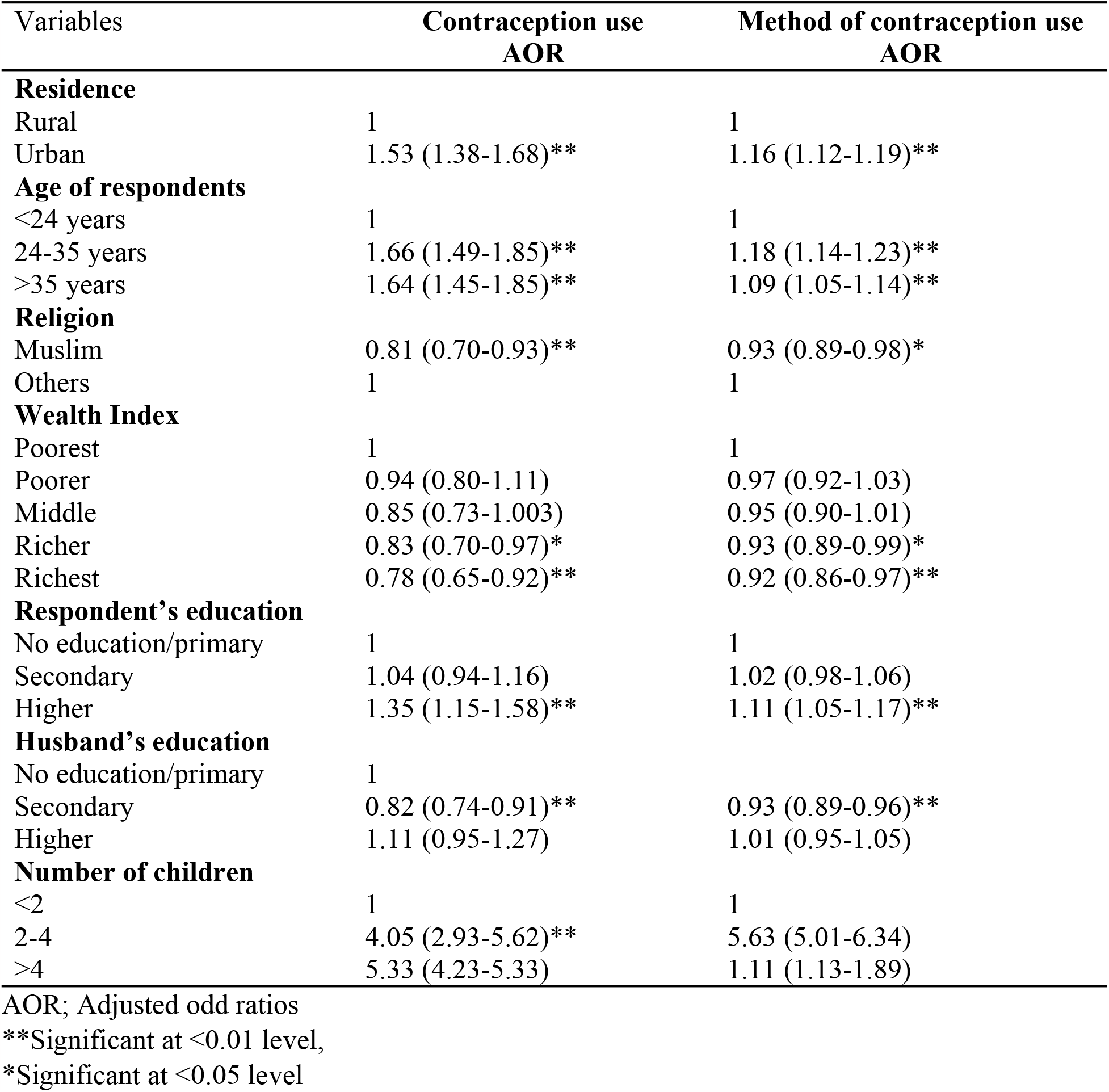
Factors affecting the use of contraception.

**Table 5:**
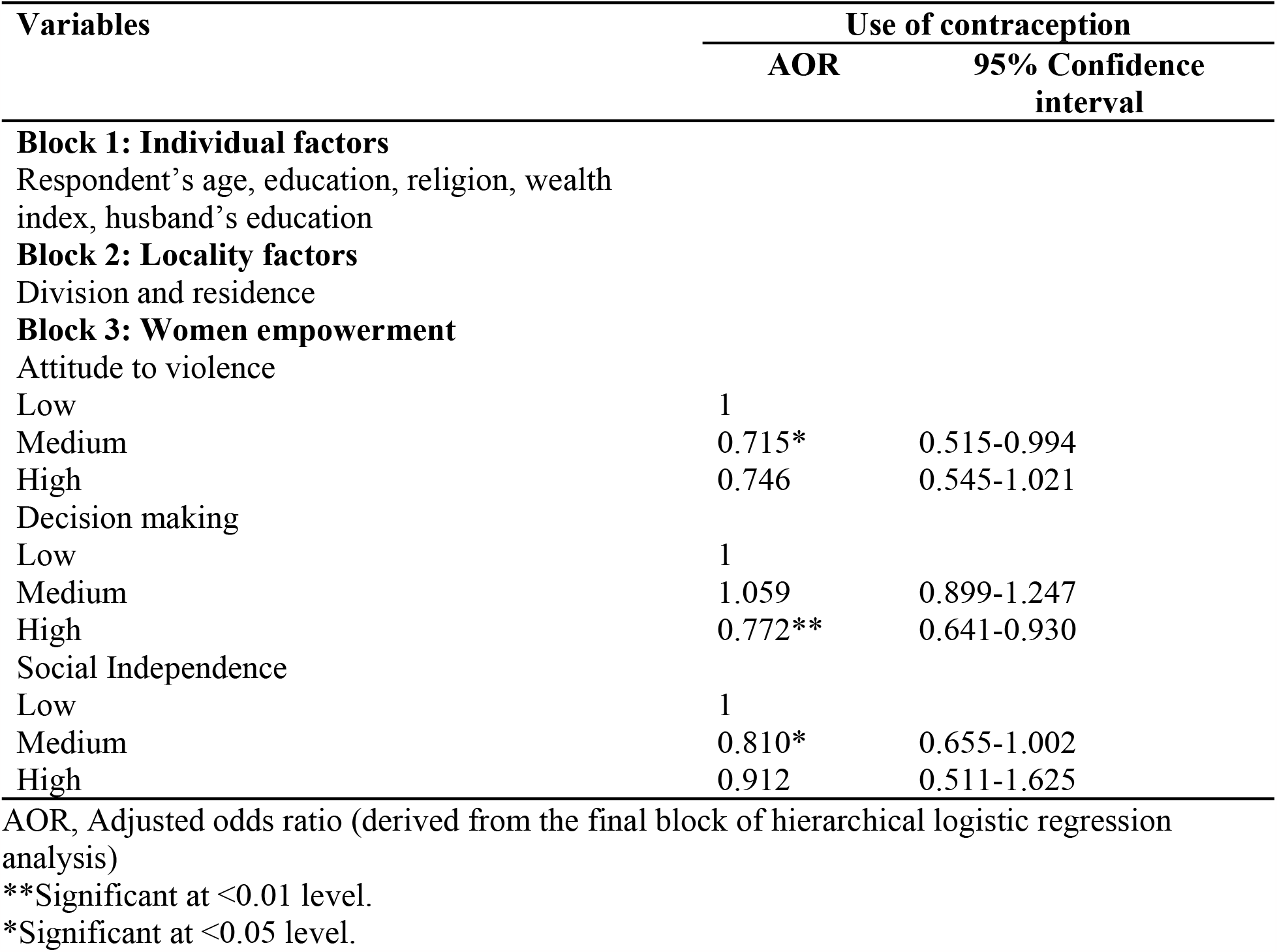
Association of women empowerment with contraception use controlling covariates factors.

| Variables | Use of contraception |  |
| --- | --- | --- |
|  | AOR | 95% Confidence interval |
| **Block 1: Individual factors** |  |  |
| Respondent's age, education, religion, wealth index, husband's education |  |  |
| **Block 2: Locality factors** |  |  |
| Division and residence |  |  |
| **Block 3: Women empowerment** |  |  |
| Attitude to violence |  |  |
| Low | 1 |  |
| Medium | 0.715* | 0.515-0.994 |
| High | 0.746 | 0.545-1.021 |
| Decision making |  |  |
| Low | 1 |  |
| Medium | 1.059 | 0.899-1.247 |
| High | 0.772** | 0.641-0.930 |
| Social Independence |  |  |
| Low | 1 |  |
| Medium | 0.810* | 0.655-1.002 |
| High | 0.912 | 0.511-1.625 |

AOR, Adjusted odds ratio (derived from the final block of hierarchical logistic regression analysis)
\*\*Significant at <0.01 level.
\*Significant at <0.05 level.

### Structural Equation Modelling

In the SEM analysis, the demographic variables that are associated with key relationships between women empowerment and contraceptive use have been controlled in the model. The model (Figure 2) showed that the value of Goodness of fit (GFI) = 0.999, CFI = 0.995 and RMSEA = 0.017 which are considered as good fit model (20, 21). The study assessed the effect of women empowerment in terms of social independence, attitude to violence and decision making on contraception use. Controlling demographic data (no. of child, women and husband’s education and women’s age), the effect of overall women empowerment on contraception use was negative and non-significant (β= -0.138, *p* = 0.193) (Figure 2).

**Figure 2:**
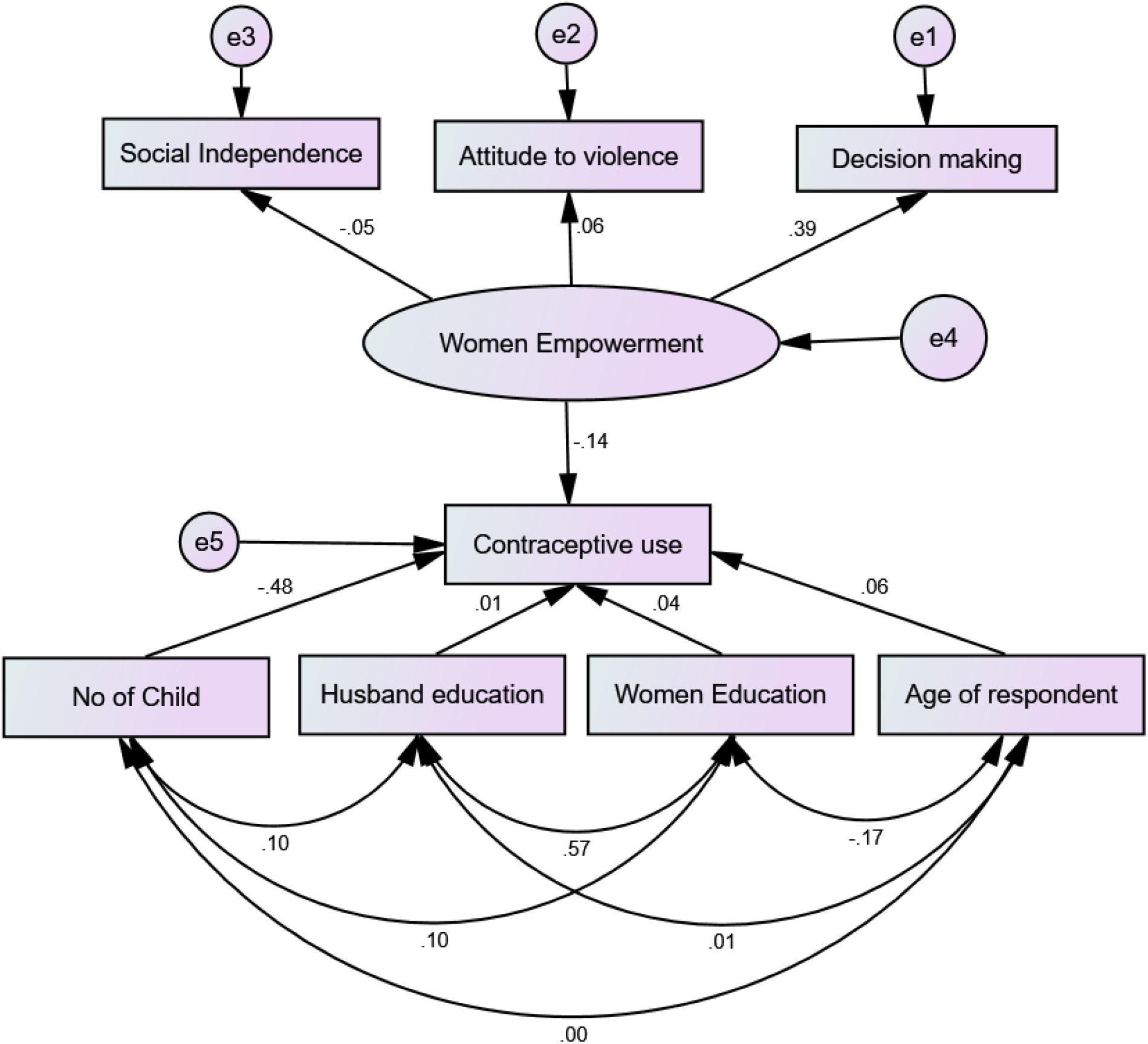
Theoretical full structural model

## Discussion

For the betterment of national demographical outcomes, women’s rights and issues in terms of women empowerment have been identified as a major contributor. This present study addressed all the aspects of women empowerment and its relationship with contraception use as well as the factors that affecting the women’s use of contraception and different methods of contraception. This paper is unique as it assesses different aspects of women empowerment through a theoretical structural model.

This study indicated a positive correlation between age and contraceptive use intervals, with older women more likely to use oral contraceptives, which is consistent with studies that were conducted in Ethiopia (22) and India (23). This mirrors a general trend, which is that women of a certain age are more motivated to engage in family planning and better comprehend the repercussions of doing so. With the increase of age, number of living children, betterment of socio-economic condition and women literacy, women decision making power increases (24-26). The findings of this study match with the findings of the National Family Health Survey-4 (23) in India, which showed that education plays in assisting women to receive knowledge about contraceptives so that they can make decisions that are based on accurate information (23).

A low rate of contraceptive use among Muslim women found in this study which are in line with those found in Rwanda (27). According to Habyarimana and Ramroop (2018) (27), this underscores the significance of taking a culturally sensitive approach when promoting, respecting, and addressing issues related to the influence of religion and cultural beliefs on family structure. The relationship between family wealth and an increased usage of contraceptives lends credence to the idea that financial resources play an essential role, which is in line with the findings of earlier research (13, 27).

The focus of the study on empowerment factors, such as aggressive intentions, decision-making power, and social autonomy, includes research carried out in India, which reveals that empowerment is a powerful factor in increasing contraceptive use among women (28-30). The findings indicate favorable benefits, such as moderate empowerment on contraceptive use, and the relevance of women’s decision-making capacity in family planning (28, 29). Similar to the findings of a study that was carried out in Nigeria (30), this study revealed that enabling factors do not appear to impact whether women use contraception medication has a role, but also contraceptive techniques (30). techniques of contraception: This study demonstrated that enabling factors do not appear to affect whether women use contraception. This shows how critically important it is for family planning initiatives to take into consideration the empowerment of women.

The results of this study demonstrate the multifaceted nature of the factors that influence women’s usage of contraceptives. These findings are backed by a large body of previous research. Continue to have a substantial impact on contraceptive use are socio-demographic factors such as age, level of education, religious affiliation, and socioeconomic standing. In addition, women’s empowerment, as assessed by their attitudes, decision-making power, and social independence, plays a vital role in promoting the use of contraception and even in shaping the form of contraception that is selected. This empowerment is measured by women’s social independence, decision-making capacity, and views.

In order for initiatives to be successful in promoting family planning and the use of contraceptives, socio-demographic and empowerment-related characteristics should be taken into consideration. According to Wado et al. 2019 (22) and Dhak et al. 2019 (23), educational efforts that target younger women and women with lower levels of education are particularly effective at raising awareness and knowledge about contraceptives. According to Habyarimana and Ramroop (2018) (27), it is essential to employ tactics that are culturally sensitive in order to address the influence of religion and cultural beliefs on decisions on family planning (28-30). Research has shown that empowerment initiatives that promote women’s participation in decision-making processes and fight against gender-based violence can contribute to higher contraceptive use and improved reproductive health outcomes. According to Agha et al. (2021) (31), increasing the use of contemporary contraceptives also requires an understanding of social norms and an effort to change those norms.

### Strengths and limitations

Our study has three main strengths. The first is use of multidimensional SWPER index to find out the relationship between women empowerment and contraception use while other studies used few components of empowerment. Secondly, we used the SEM analysis which helps us to explain the relationship as well as to examine the direct and indirect relationships of the variables. The third one is that we used nationally representative data covering both rural and urban regions and also had large sample size that reduced the biasness in the study.

The study is not without limitations. Although the study used multidimensional SWPER index for estimating women empowerment, it is difficult to capture all the domains of women empowerment in a single index. As the study used secondary data, there are some limitations in data analysis due to available variables. We used data of women who are at reproductive age and married which should not be generalized for women of all ages. Finally, the study can not conclude causal-effect relationship due to being cross sectional study.

## Conclusion

This research highlights the significance of women’s empowerment in influencing contraceptive use in Bangladesh. The study emphasizes that women empowerment alone cannot influence the use of contraception. Age, education, and socioeconomic level are all important predictors of contraceptive use. Cultural and religious influences, especially among Muslim women, emphasize the importance of culturally sensitive approaches to family planning activities. Women’s decision-making capacity, social autonomy and husband’s involvement are important enabling variables in increasing contraceptive use. Initiatives that promote women’s participation in decision-making and reduce gender-based violence can improve reproductive health outcomes. Tailored interventions that take socioeconomic and empowering variables into account are critical for promoting family planning and reproductive health in Bangladesh. More research is required to investigate these processes in greater depth and develop targeted tactics.

## Acknowledgments

Authors are thankful to all the principal investigators of the Demographic Health Survey for their relentless efforts in producing such vital information for community-based estimates.

## Data availability

Data generated for analysis of the present study will be made available upon request.

## References

1. Malhotra A, Schuler SR, Boender C, editors. Measuring women’s empowerment as a variable in international development. background paper prepared for the World Bank Workshop on Poverty and Gender: New Perspectives; 2002: The World Bank Washington, DC.

2. Klima aKlimaS. Unintended pregnancy: consequences and solutions for a worldwide problem. Journal of Nurse-Midwifery. 1998;43(6):483–91.

3. Victora CG, Requejo JH, Barros AJ, Berman P, Bhutta Z, Boerma T, et al. Countdown to 2015: a decade of tracking progress for maternal, newborn, and child survival. The Lancet. 2016;387(10032):2049–59.

4. Germain A, Kyte R. The Cairo consensus: the right agenda for the right time: International Women’s Health Coalition New York, NY; 1995.

5. Schuler SR, Hashemi SM, Riley RileyP. The influence of women’s changing roles and status in Bangladesh’s fertility transition: evidence from a study of credit programs and contraceptive use. World Development. 1997;25(4):563–75.

6. Blanc aBlancK. The effect of power in sexual relationships on sexual and reproductive health: an examination of the evidence. Studies in family planning. 2001;32(3):189–213.

7. Uddin J, Pulok MH, Sabah MN-U. Correlates of unmet need for contraception in Bangladesh: does couples’ concordance in household decision making matter? Contraception. 2016;94(1):18–26.

8. National Institute of Population Research and Training (NIPORT) II. Bangladesh Demographic and Health Survey 2017–18.. Dhaka, Bangladesh.; 2020.

9. Lasee A, Becker S. Husband-wife communication about family planning and contraceptive use in Kenya. International family planning perspectives. 1997:15–33.

10. Bawah aBawahA. Spousal communication and family planning behavior in Navrongo: a longitudinal assessment. Studies in family planning. 2002;33(2):185–94.

11. Bogale B, Wondafrash M, Tilahun T, Girma E. Married women’s decision making power on modern contraceptive use in urban and rural southern Ethiopia. BMC public health. 2011;11(1):1–7.

12. Economic UNDoI, Economic UNDf, Information S, Analysis P. World population prospects: United Nations, Department of International, Economic and Social Affairs; 1984.

13. Biswas TK, Kabir M. Women’s Empowerment and Current use of Contraception in Bangladesh. Asia-Pacific Journal of Rural Development. 2002;12(2):1–13.

14. Hasan MS, Dana GPT, Jafrin N. Determinants of contraceptive use in rural Bangladesh. Journal of Population and Development. 2014;1:53–72.

15. Hameed W, Azmat SK, Ali M, Sheikh MI, Abbas G, Temmerman M, et al. Women’s empowerment and contraceptive use: the role of independent versus couples’ decision-making, from a lower middle income country perspective. PloS one. 2014;9(8):e104633.

16. Shariful Islam S, Mainuddin A. Relationship between income generating activities of rural women and their reproductive health behavior in Bangladesh. Rural and Remote Health. 2015;15(4):53–63.

17. Islam aIslamZ. Factors affecting modern contraceptive use among fecund young women in Bangladesh: does couples’ joint participation in household decision making matter? Reproductive health. 2018;15(1):1–9.

18. Ewerling F, Raj A, Victora CG, Hellwig F, Coll CV, Barros BarrosJ. SWPER Global: A survey-based women’s empowerment index expanded from Africa to all low-and middle-income countries. Journal of global health. 2020;10(2).

19. Ewerling F, Lynch JW, Victora CG, van Eerdewijk A, Tyszler M, Barros BarrosJ. The SWPER index for women’s empowerment in Africa: development and validation of an index based on survey data. The Lancet Global Health. 2017;5(9):e916–e23.

20. Hatcher L, O’Rourke N. A step-by-step approach to using SAS for factor analysis and structural equation modeling: Sas Institute; 2013.

21. Hu Lt, Bentler BentlerM. Cutoff criteria for fit indexes in covariance structure analysis: Conventional criteria versus new alternatives. Structural equation modeling: a multidisciplinary journal. 1999;6(1):1–55.

22. Wado YD, Gurmu E, Tilahun T, Bangha M. Contextual influences on the choice of long-acting reversible and permanent contraception in Ethiopia: a multilevel analysis. PloS one. 2019;14(1):e0209602.

23. Dhak B, Saggurti N, Ram F. Contraceptive use and its effect on Indian women’s empowerment: evidence from the National Family Health Survey-4. Journal of Biosocial Science. 2020;52(4):523–33.

24. Haque M, Islam TM, Tareque MI, Mostofa M. Women empowerment or autonomy: A comparative view in Bangladesh context. Bangladesh e-journal of Sociology. 2011;8(2):17–30.

25. Acharya DR, Bell JS, Simkhada P, Van Teijlingen ER, Regmi RegmiR. Women’s autonomy in household decision-making: a demographic study in Nepal. Reproductive health. 2010;7(1):1–12.

26. Saleem S, Bobak M. Women’s autonomy, education and contraception use in Pakistan: a national study. Reproductive health. 2005;2(1):1–8.

27. Habyarimana F, Ramroop S. Spatial analysis of socio-economic and demographic factors associated with contraceptive use among women of childbearing age in Rwanda. International journal of environmental research and public health. 2018;15(11):2383.

28. León FR, Lundgren R, Sinai I, Sinha R, Jennings V. Increasing literate and illiterate women’s met need for contraception via empowerment: a quasi-experiment in rural India. Reproductive health. 2014;11:1–10.

29. Nadeem M, Malik MI, Anwar M, Khurram S. Women decision making autonomy as a facilitating factor for contraceptive use for family planning in Pakistan. Social Indicators Research. 2021;156:71–89.

30. OlaOlorun FM, Anglewicz P, Moreau C. From non-use to covert and overt use of contraception: Identifying community and individual factors informing Nigerian women’s degree of contraceptive empowerment. PloS one. 2020;15(11):e0242345.

31. Agha S, Morgan B, Archer H, Paul S, Babigumira JB, Guthrie GuthrieL. Understanding how social norms affect modern contraceptive use. BMC Public Health. 2021;21(1):1–11.

